# Autoimmune responses to myelin-associated proteins as diagnostic and prognostic biomarkers of relapsing-remitting multiple sclerosis: associations with human herpesvirus-6 and Epstein-Barr Virus reactivation

**DOI:** 10.1101/2024.11.16.24317434

**Authors:** Aristo Vojdani, Abbas F. Almulla, Elroy Vojdani, Jing Li, Yingqian Zhang, Michael Maes

## Abstract

**Background:** The pathogenesis of relapsing-remitting multiple sclerosis (RRMS) is linked to autoimmune attacks against myelin proteins, and reactivation of Epstein-Barr virus (EBV) and human herpesvirus 6 (HHV-6). However, the connection between viral reactivation and autoimmune biomarkers has remained unclear.

**Objectives:** To investigate immunoglobulin (Ig)G/IgA/IgM responses targeting myelin-related proteins in association with EBV and HHV-6 replication markers in RRMS.

**Methods:** We recruited 55 patients with RRMS and 63 healthy controls and assessed IgG/IgA/IgM responses against seven myelin-related components, as well as EBV nuclear antigen 1 (EBNA-1) and deoxyuridine-triphosphate nucleotidohydrolase (dUTPases). Disability was evaluated using the Expanded Disability Status Scale (EDSS) and disease progression using the Multiple Sclerosis Severity Score (MSSS).

**Results:** IgG/IgA/IgM levels targeting seven myelin-related proteins were significantly higher in RRMS than in controls. IgG against myelin basic protein (MBP) (IgG-MBP), IgM-myelin-associated glycoprotein (IgM-MAG)-37-60, IgA-MBP, and IgA-myelin-oligodendrocyte-glycoprotein (IgA-MOG-31-55) distinguished RRMS from controls with a predictive accuracy of 96.6% (sensitivity = 95.7%, specificity = 95.2%) and an area under the ROC curve of 0.991. A large part of the variance in the EDSS (around 75%) and MSSS score (62.8%) was explained by IgG-MBP, IgM-MBP, IgA-MOG-31-55, and IgM-MAG. Part of the variance (47.4%) in the IgG/IgA/IgM responses to myelin-related proteins was explained by immune responses to EBNA and deoxyuridine-triphosphate nucleotidohydrolases of EBV and HHV-6.

**Conclusions:** Autoimmune reactivities targeting myelin-related proteins are valuable biomarkers of RRMS and the severity and progression of RRMS. Reactivation of EBV and HHV-6 may trigger or maintain these autoimmune responses thereby impacting disease progression.

## Introduction

Multiple sclerosis (MS) is a chronic, immune-mediated disorder of the central nervous system (CNS) characterized by inflammatory demyelination and progressive neurological damage [1]. Among its subtypes, relapsing-remitting MS (RRMS) is the most prevalent, comprising 63% to 80% of MS cases globally [2]. RRMS features episodes of acute relapses, followed by periods of remission where symptoms may improve partially or fully [3]. These relapses coincide with CNS inflammation and demyelination, while remission reflects an incomplete repair process [4]. The fluctuating nature of RRMS can obscure underlying progressive damage, as even during remissions, inflammatory processes persist and contribute to long-term neurodegeneration [5, 6].

The onset and progression of RRMS are influenced by genetic and environmental factors [7, 8]; for example, the presence of HLA-DRB1*15:01 allele, lowered vitamin D, smoking and viral infections were all reported as significant trigger factors of MS [9–12]. Moreover, activated immune inflammatory system (IRS) responses, as indicated by increased T helper (Th)1 and Th17 activities, and a diminished compensatory immune-regulatory response (CIRS), as indicated by decreased Th2 activity [13, 14], along with increased oxidative and nitrosative stress are also implicated [8, 15]. Breakdown of immune tolerance may cause autoreactive T cells and B cells to attack CNS proteins such as myelin basic protein (MBP), proteolipid protein (PLP), myelin oligodendrocyte glycoprotein (MOG), and oligodendrocytic basic protein [16–18]. These proteins are essential for maintaining myelin integrity, and their disruption leads to the hallmark demyelination seen in RRMS [19]. The presence of elevated autoantibodies targeting myelin proteins may serve as valuable diagnostic and prognostic markers, correlating with increased disease severity and disability in RRMS patients [20]. However, there is no data indicating which of the IgG, IgA, or IgM responses to those antigens is most effective in differentiating RRMS from controls, nor which myelin-associated proteins exhibit the highest predictive accuracy.

Autoreactive T cells, particularly Th1 and Th17 subsets, may infiltrate the CNS and initiate inflammatory attacks on myelin sheaths, leading to demyelination and neurodegeneration (Dendrou et al., 2015). B cells also contribute significantly by producing autoantibodies, presenting antigens, and stimulating the secretion of pro-inflammatory cytokines (Getahun and Cambier, 2019). The delivery of these immune mediators into the CNS further triggers microglia and astrocytes, which further exacerbate the immune response by releasing neurotoxic cytokines and reactive oxygen species, leading to progressive axonal loss and disabilities (Kunkl et al., 2022). However, the precise triggers of this immune dysregulation in RRMS are still not fully understood.

A growing body of evidence points to viral reactivation, particularly Epstein-Barr virus (EBV) and Human Herpesvirus 6 (HHV-6), as a key driver of the immune dysfunctions in RRMS [21, 22]. Recently, high levels of immunoglobulins (Ig)A/IgG/IgM targeting deoxyuridine-triphosphate nucleotidohydrolase (dUTPase) of HHV-6 and EBV, and EBV nuclear antigen (EBNA) were detected in RRMS, indicating reactivation of both viruses [23]. Moreover, the increased reactivities against those viral antigens were associated with abnormal M1 macrophage and Th17 activities along with overall IRS activation [23]. Reactivation of these latent viruses was found to be associated with disabilities, higher severity, and neuropsychiatric symptoms in RRMS [23, 24]. Increased antibodies contribute to CNS damage through processes like complement activation and antibody-dependent cell-mediated cytotoxicity (Phares et al., 2011). Additionally, viral reactivation may disrupt immune regulation by triggering bystander activation, where non-specific immune cells become activated and exacerbate inflammation (Fujinami et al., 2006). However, the associations between HHV-6 and EBV replication and autoimmune responses to myelin-related proteins remain poorly understood.

Hence, this study aims to evaluate the IgA, IgM, and IgG immune responses to key myelin-related proteins, including MBP, PLP, MOG-35-55, MOG-31-55, citrullinated-MOG (CIT-MOG), myelin-associated glycoprotein (MAG)-37-60, and GLIAL-CAM-370-399, in RRMS patients. We explore the diagnostic performance of these autoimmune responses for RRMS, and which proteins show the best predictive accuracy. In addition, we examine their associations with severity of RRMS and disease progression. Finally, the study will investigate the associations between these immune responses and the reactivation of latent viruses such as EBV and HHV-6.

## Subjects and Methods

### Subjects

This case-control study involved the recruitment of 55 patients diagnosed with RRMS from the Neuroscience Centre at Alsader Medical City in Al-Najaf, Iraq. All participants were in the remission phase of the condition. The diagnosis was established according to the McDonald criteria [25] and confirmed by a senior neurologist with significant expertise in multiple sclerosis. A control group comprising 63 healthy individuals from the same geographic region, including hospital staff, their acquaintances, and associates of MS patients, was selected for comparative analysis.

Patients and controls underwent comprehensive screening to eliminate individuals with a prior lifetime diagnosis of Axis I neuropsychiatric disorders, as outlined by the DSM-IV-TR. The conditions included major depressive disorder (MDD), generalized anxiety disorder, obsessive-compulsive disorder, panic disorder, psycho-organic conditions unrelated to MS, schizophrenia, post-traumatic stress disorder, substance use disorders (excluding nicotine dependence), bipolar disorder, and autism spectrum disorders. Additionally, individuals with a history of medical conditions, including thyroid disorders, renal or liver disease, type 1 diabetes mellitus, cardiovascular diseases, or (auto)immune disorders such as cancer, inflammatory bowel disease, rheumatoid arthritis, chronic obstructive pulmonary disease, psoriasis, lupus erythematosus, and chronic fatigue syndrome/myalgic encephalomyelitis (CFS/ME) were systematically excluded from the study. Furthermore, individuals with neurodegenerative diseases, such as Parkinson’s and Alzheimer’s disease, as well as control subjects, were excluded from the study.

Informed consent was obtained and documented from all RRMS patients, or their legal guardians or parents when applicable, as well as from control subjects prior to enrolment in the study. The research obtained ethical approval from the institutional ethics committee of the College of Medical Technology at The Islamic University of Najaf, Iraq, as indicated in document reference 11/2021. The study complied with ethical standards established by national and international frameworks, including the Declaration of Helsinki by the World Medical Association, The Belmont Report, CIOMS guidelines, and the International Conference on Harmonization of Good Clinical Practice (ICH-GCP). The Institutional Review Board (IRB) ensured compliance with international guidelines to protect the well-being of human research participants.

### Clinical Assessments

The senior neurologist performed a semi-structured interview to collect critical sociodemographic and clinical information. The evaluation of clinical disabilities was conducted utilizing the Expanded Disability Status Scale (EDSS), which was initially developed by Kurtzke in 1983 [26]. The Multiple Sclerosis Severity Score (MSSS), created by Roxburgh, Seaman, and colleagues in 2005, was employed to assess the rate of disability progression [27]. Furthermore, the body mass index (BMI) of each participant was calculated by dividing their weight in kilograms by the square of their height in meters.

### Biomarkers assays

Fasting blood samples were obtained via venipuncture, using disposable equipment, from participants between 7:30 and 9:00 a.m. The samples were left to coagulate at room temperature for 15 minutes before being centrifuged at 3500 RPM for 10 minutes. The resulting serum was divided into Eppendorf tubes for various assay purposes. MBP was obtained from Sigma-Aldrich (St. Louis, MO USA). PLP, MOG-31-55, MOG-35-55, CIT MOG-31-55, MAG-37-60, GLIAL-CAM-370-399 and peptides specific to HHV-6-dUTPase, EBV-dUTPase, and EBNA-366-406 (EBNA) were synthesized by Biosynthesis (Lewisville, TX USA). These antigens and peptides were used in enzyme-linked immunosorbent assays (ELISA). Detailed descriptions of these methods can be found in prior publications [28, 29]. In summary, ELISA plates were coated with 100 microliters of each peptide, at an optimal concentration of 5-10 µg/mL in 0.1 M carbonate buffer (pH 9.5). After incubation, plates were washed and blocked with 2% BSA. Following that, 100 microliters of serum from MS patients and healthy controls, diluted to 1:50 for IgA and 1:100 for IgG and IgM, were added to the wells in duplicate and incubated at 25° for one hour. Plates were again washed and incubated before secondary antibodies were introduced. After repeated washing and substrate addition, color development was measured, and indices were calculated using standard sera from MS patients as calibrators and controls. The study analyzed ten biomarkers, which included IgG, IgA, and IgM responses to EBNA, as well as EBV and HHV-6-dUTPase peptides.

### Data analysis

All statistical analyses in this research were conducted using IBM SPSS version 29. The comparison of continuous variables across diverse groups was performed using analysis of variance (ANOVA), while contingency table analysis was employed to assess associations between categorical variables. The False Discovery Rate (FDR) p-correction was used to adjust for multiple comparisons. Pearson’s product-moment correlation was used to explore connections among scale variables. Multivariate regression analysis was carried out to evaluate how autoimmune reactivity to CNS proteins influenced disability and symptom severity scores. Adjustments were made for demographic factors such as age, gender, BMI, and smoking habits. Both manual and stepwise methods were utilized, with inclusion and exclusion criteria set at p-values of 0.05 and 0.06, respectively. Several key parameters, including standardized beta coefficients, degrees of freedom (df), p-values, R², and F-statistics, were reported in the analysis. To assess heteroskedasticity, the White test and a modified Breusch-Pagan test were used, while collinearity was evaluated based on tolerance and variance inflation factor (VIF). All tests employed a two-tailed approach with a significant threshold of 0.05.

Principal component analysis (PCA) was used to reduce features and assess the extraction of a single “general” principal component from variables, such as severity ratings, disability scores, and viral antigens. The PCA was considered valid when the Kaiser-Meyer-Olkin (KMO) value exceeded 0.6, the first principal component explained more than 50% of the variance, and all loadings on this component surpassed 0.7. Canonical correlation analysis (CCA) was employed to investigate the relationships between two sets of variables: autoantibodies against CNS proteins as the dependent set, and autoantibodies against viral antigens as the explanatory set. Canonical variables were considered valid if the explained variance exceeded 50% and all canonical loadings were greater than 0.5. Additionally, we assessed the proportion of variance in the clinical data that could be explained by the biomarker data.

Furthermore, multilayer perceptron neural networks were developed to differentiate RRMS patients from healthy controls. These models used IgG, IgM, and IgA responses to CNS proteins, such as MBP, MOG, and MAG, as input variables. The neural networks were designed with a feedforward architecture, consisting of two hidden layers, each containing up to eight nodes, and were trained in batch mode for up to 250 epochs. Training ceased when there were no further reductions in the error term. The importance of input variables was visualized using importance charts, and performance metrics, including error rates, relative errors, and misclassification rates, were computed by comparing predicted versus actual values.

A preliminary power analysis using G*Power 3.1.9.7 determined that a minimum sample size of 73 participants was necessary to assess the deviation of R² from zero in a fixed-model linear multiple regression analysis. This calculation was based on a power of 0.8, a significance level of 0.05, four covariates, and an effect size of 0.176.

## Results

### Sociodemographic and Clinical Features of MS

Electronic supplementary file (ESF), Table 1 summarizes the sociodemographic details and disability assessment scores, namely EDSS and MSSS, comparing patients with RRMS to healthy controls. No significant differences were found between the two groups regarding age, gender, marital status, or BMI. However, notable distinctions were observed in employment and smoking status. Additionally, the RRMS group had substantially higher EDSS and MSSS scores, indicating more severe disabilities. The PC scores of IgA, IgM, and IgG antibodies directed against dUTPase-EBV, dUTPase-HHV-6, and EBNA were also elevated in RRMS patients compared to the control group. The latter were computed as the first PCs extracted from all IgM, IgG, and IgA responses to EBNA, EBV dUTPase or HHV-6 dUTPase (labeled PC_3EBNAs, PC_3dUTPases-EBV, and PC_3dUTPases-HHV-6). As such, these PCs combine the three immunoglobulin responses directed against the three viral antigens.

**Table 1.** Associations among autoimmune biomarkers and relapsing-remitting multiple sclerosis (RRMS) versus healthy controls (HC).

| **Variables** | **Healthy Controls**<br>**(n=63)** | **RRMS Patients**<br>**(n=55)** | **F/$\chi^2$** | **df** | **p-value** |
| --- | --- | --- | --- | --- | --- |
| IgG-MBP | 0.852(0.063) | 2.267(0.067) | 218.05 | 1/112 | <0.001 |
| IgG-MBP | 0.632(0.055) | 1.270(0.060) | 56.52 | 1/112 | <0.001 |
| IgA-MBP | 0.563(0.055) | 1.274(0.059) | 71.83 | 1/112 | <0.001 |
| IgG-PLP | 0.744(0.063) | 1.092(0.067) | 13.16 | 1/112 | <0.001 |
| IgM-PLP | 0.481(0.038) | 0.744(0.040) | 20.74 | 1/112 | <0.001 |
| IgA-PLP | 0.419(0.046) | 0.680(0.049) | 13.90 | 1/112 | <0.001 |
| IgG-MOG-35-55 | 0.365(0.035) | 0.591(0.037) | 18.11 | 1/112 | <0.001 |
| IgM-MOG-35-55 | 0.263(0.022) | 0.484(0.024) | 41.40 | 1/112 | <0.001 |
| IgA-MOG-35-55 | 0.258(0.021) | 0.438(0.023) | 31.38 | 1/112 | <0.001 |
| IgG-MOG-31-55 | 0.782(0.085) | 1.715(0.092) | 51.12 | 1/112 | <0.001 |
| IgM-MOG-31-55 | 0.378(0.044) | 0.861(0.048) | 50.66 | 1/112 | <0.001 |
| IgA-MOG-31-55 | 0.376(0.049) | 1.038(0.053) | 78.14 | 1/112 | <0.001 |
| IgG-CIT-MOG | 0.478(0.037) | 0.870(0.040) | 47.02 | 1/112 | <0.001 |
| IgM-CIT-MOG | 0.295(0.029) | 0.624(0.031) | 53.99 | 1/112 | <0.001 |
| IgA-CIT-MOG | 0.387(0.037) | 0.640(0.040) | 19.98 | 1/112 | <0.001 |
| IgG-MAG-37-60 | 0.705(0.093) | 1.584(0.100) | 38.33 | 1/112 | <0.001 |
| IgM-MAG-37-60 | 0.468(0.063) | 1.259(0.068) | 66.16 | 1/112 | <0.001 |
| IgA-MAG-37-60 | 0.434(0.051) | 0.952(0.055) | 44.44 | 1/112 | <0.001 |
| IgG-GLIAL-CAM-370-399 | 0.468(0.096) | 1.249(0.103) | 28.27 | 1/112 | <0.001 |
| IgM-GLIAL-CAM-370-399 | 0.325(0.028) | 0.583(0.030) | 36.81 | 1/112 | <0.001 |
| IgA-GLIAL-CAM-370-399 | 0.269(0.027) | 0.565(0.029) | 52.05 | 1/112 | <0.001 |

All results of univariate General Linear Models. All data are shown as marginal estimated mean (SE) values after covarying for age, sex, body mass index, and smoking.
Ig: Immunoglobulin, MBP: myelin basic protein, PLP: myelin basic protein-proteolipid protein complex, MOG-35-55: myelin oligodendrocyte glycoprotein- amino acids 35 to 55, MOG-31-55: myelin oligodendrocyte glycoprotein- amino acids 31 to 55, CIT-MOG: citrullinated myelin oligodendrocyte glycoprotein, MAG-37-60: myelin-associated glycoprotein-amino acids 37 to 60.

### Differences in Autoimmune Responses to CNS Proteins Among Study Groups

**Table 1** demonstrates that RRMS patients showed significantly higher IgG, IgA, and IgM responses to MBP, PLP, MOG-35-55, MOG-31-55, CIT-MOG, MAG-37-60, and GLIAL-CAM-370-399, compared to the healthy control group. The highest effect sizes, as indicated by Partial Eta Squared, were in descending order: IgG-MBP (0.661), IgA-MOG-31-55 (0.411), IgA-MBP (0.391), and IgM-MAG-37-60 (0.371). **Figure 1** shows the scatter plot of these 4 biomarkers in RRMS *versus* controls.

**Figure 1.**
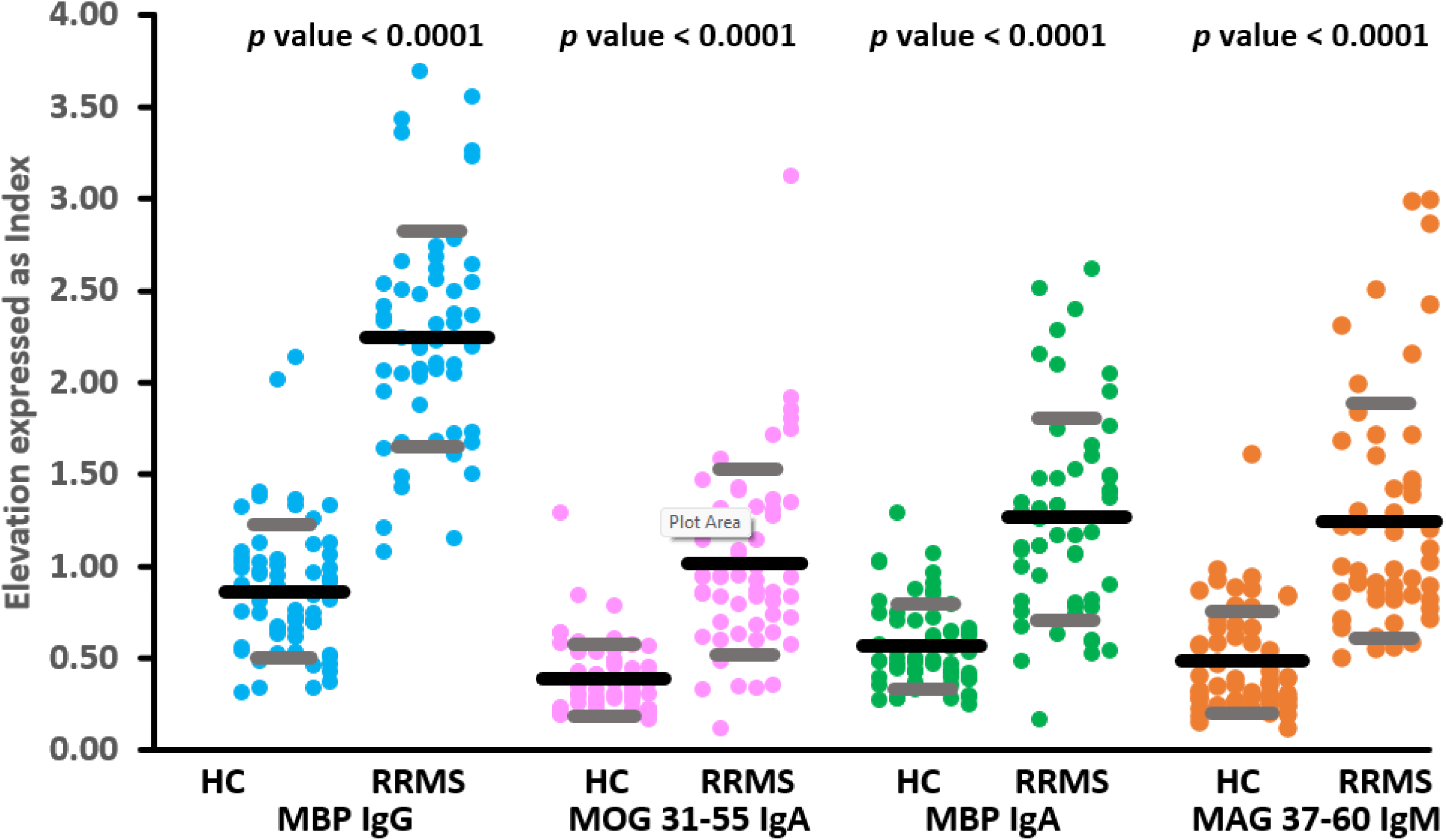
Dot-scatter plot (mean and standard deviation) of the top 4 autoantibody levels to myelin-related proteins distinguishing relapsing-remitting multiple sclerosis (RRMS) from healthy controls (HC). MBP IgG: IgG against myelin basic protein MOG-31-55 IgA: IgA directed to myelin-oligodendrocyte-glycoprotein MBP IgA: IgA directed against MBP MAG-IgM: IgM directed to myelin-associated glycoprotein.

### Diagnostic performance

We conducted a binary logistic regression to identify the most significant predictors of RRMS. In this analysis, the control group served as the reference, with RRMS as the dependent variable. Logistic regression showed that the best discrimination of RRMS from controls was obtained when IgG-MBP was combined with IgM-MAG-37-60. Model #1 from **Table 2** showed that both IgG-MBP and IgM-MAG-37-60 were significantly associated with RRMS, with an effect size of 0.922 and an overall accuracy of 96.6% (sensitivity = 96.4%, specificity = 96.8%).

**Table 2:** Results of binary logistic regression analysis with relapsing-remitting multiple sclerosis (RRMS) as dependent variable and healthy controls as reference group.

| Dichotomies |  | Explanatory Variables | B | SE | Wald | P | OR | 95% CI |
| --- | --- | --- | --- | --- | --- | --- | --- | --- |
| RRMS<br>versus healthy<br>controls | Model<br>#1 | IgG-MBP | 5.15 | 1.277 | 16.27 | <0.001 | 172.68 | 14.13; 2111.10 |
|  |  | IgM-MAG-37-60 | 3.80 | 1.297 | 8.61 | 0.003 | 44.98 | 3.54; 571.73 |

Ig: Immunoglobulin, MBP: myelin basic protein, MAG-37-60: myelin-associated glycoprotein-amino acids 37 to 60 of the protein.

**Table 3** presents the results of neural network analysis. Neural network model NN#1, designed to differentiate RRMS patients from healthy controls, consisted of two hidden layers: 4 units in the first layer, 3 in the second, and 2 units in the output layer. The error term in the testing sample was lower than in the training sample, while the error rate remained consistent across the three datasets, indicating no overtraining. This model performed better than logistic regression, achieving 100% accuracy in the holdout sample (sensitivity = 100%, specificity = 100%) with an ROC area of 0.989. The key biomarkers, ranked by importance, were IgG-MBP and IgM-MAG-37-60, as shown in **Figure 2**. Two markers marginally contributed to the prediction of the model, namely IgA-PLP and IgG-PLP. Therefore, we re-ran the neural network without these two markers (see NN#2).

**Figure 2.**
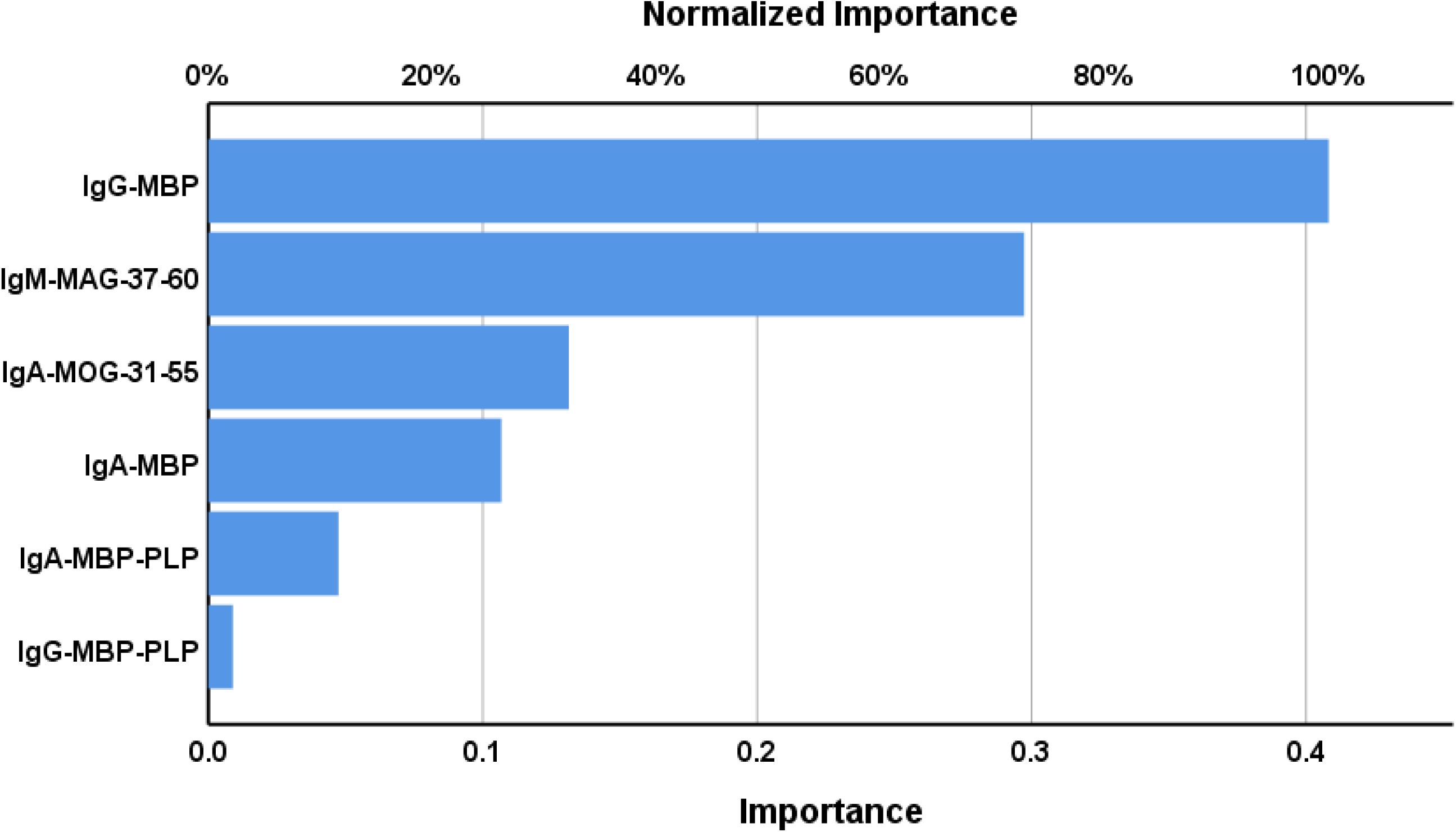
Results of neural network analysis demonstrating the importance chart. The output variables are the diagnosis of relapsing remitting multiple sclerosis and healthy controls. Input variables are, Immunoglobulin responses targeting myelin-related proteins.

**Table 3.** Results of neural networks (NN) with the diagnosis relapsing-remitting multiple sclerosis (RRMS) as output variables and autoimmune biomarkers as input data

|  | **Models** | **NN#1** | **NN#2** |
| --- | --- | --- | --- |
| **Input Layer** | Number of units | 6 | 4 |
| **Hidden layers** | Number of hidden layers | 2 | 2 |
|  | Activation function | Hyperbolic tangent | Hyperbolic tangent |
|  | Number of units in hidden layer 1 | 4 | 3 |
|  | Number of units in hidden layer 2 | 3 | 2 |
| **Output layer** | Dependent variable | RRMS Patients vs controls | RRMS Patients vs controls |
|  | Number of units | 2 | 2 |
|  | Activation function | Identity | Identity |
| **Training** | Error term | 2.775 | 3.161 |
|  | %incorrect or relative error | 7.0% | 7.7% |
|  | Prediction (sens, spec) | 91.7%, 93.9% | 95.5%, 90.0% |
| **Testing** | Sum of Squares error | 0.508 | 0.860 |
|  | % incorrect or relative error | 0% | 4.5% |
|  | Prediction: sens, spec | 100.0%, 100.0% | 100.0%, 91.7% |
|  | AUC ROC | 0.989 | 0.991 |
| **Holdout** | %incorrect or relative error | 0% | 4.5% |
|  | Predictive accuracy: sens, spec | 100.0%, 100.0% | 95.7%, 95.2% |

The NN#2 model used two hidden layers, with 3 units in the first layer, 2 in the second, and 2 units in the output layer. The error term for the testing sample was lower than for the training sample, and error rates were comparable across the training, testing, and holdout datasets. NN#2 showed a predictive accuracy of 95.5% in the holdout sample (sensitivity = 95.7%, specificity = 95.2%) and an ROC area of 0.991. **Figure 3** shows the importance chart. The most influential biomarkers were IgG-MBP and IgM-MAG-37-60, followed by IgA-MBP and IgA-MOG-31-55.

**Figure 3.**
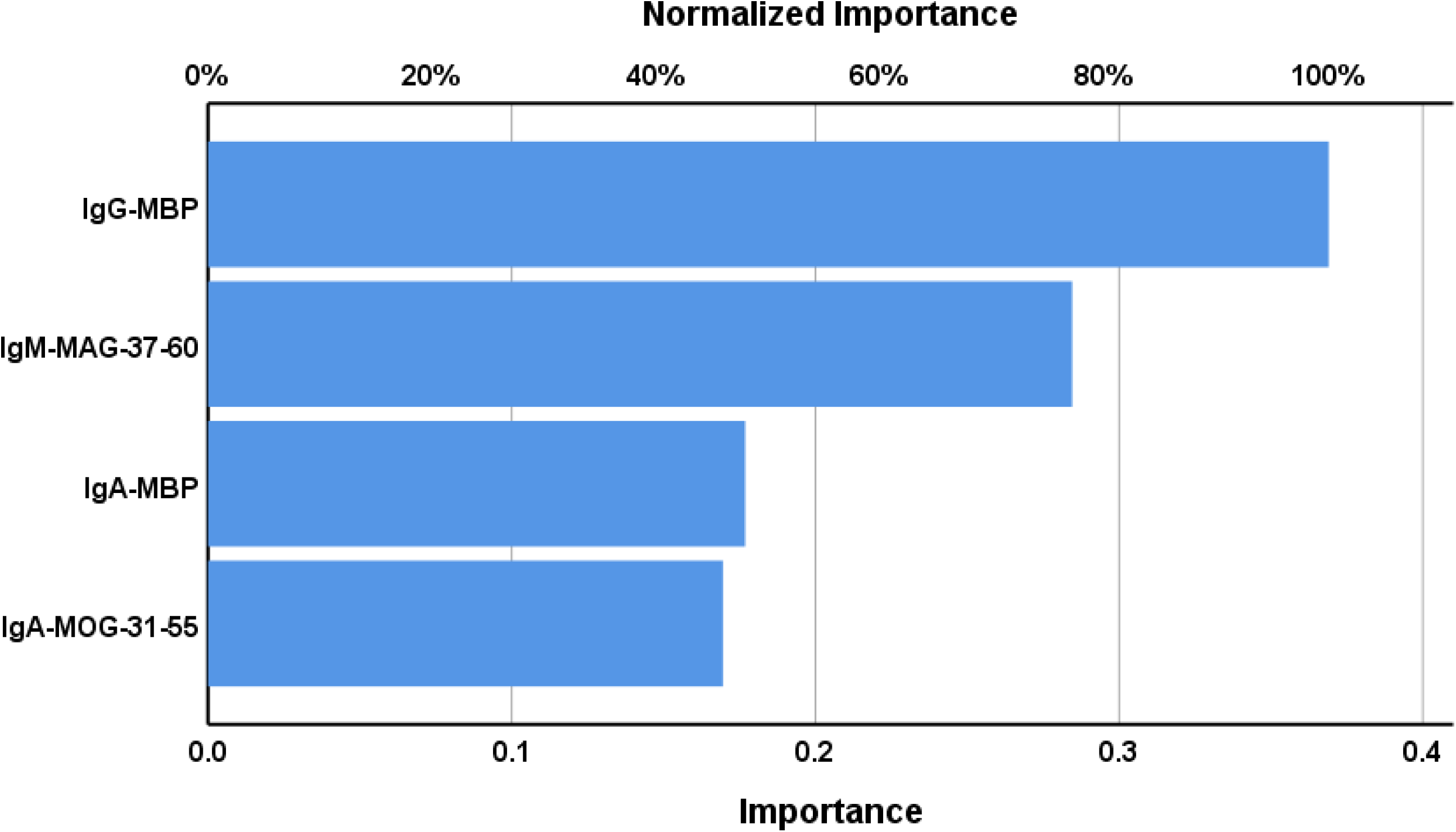
Results of neural network analysis demonstrating the importance chart. The output variables are the diagnosis of relapsing remitting multiple sclerosis and healthy controls. Input variables are, Immunoglobulin responses targeting myelin-related proteins.

### Prediction of RRMS severity by autoimmune responses to myelin-related proteins

**Table 4** details the results of multiple regression analyses, with EDSS and MSSS scores as the dependent variables. The predictors included IgG and IgM to MBP, IgA to MOG-31-55, IgM to MAG-37-60, IgA to CIT-MOG, IgA to GLIAL-CAM-370-399. We allowed for the effects of age, sex, smoking, and BMI. In the first regression, 75.6% of the variance in EDSS scores was explained by IgG-MBP, IgA-MOG-31-55, IgM-MAG-37-60, and sex. **Figure 4** illustrates the partial regression plot of the EDSS scores on IgG-MBP. The second regression model demonstrates that IgA-MOG-31-55, IgG-MBP, and IgM-MBP explained 62.8% of the variance in the MSSS score. Additional analyses were conducted (regression #3 and #4) on RRMS patients only. Regression #3 indicated that 7.4% of the variance in the EDSS score could be attributed to IgA-CIT-MOG, showing a positive association. Finally, regression #4 found that IgA-GLIAL-CAM-370-399 accounted for around 20.5% of the variance in MSSS scores. **Figure 5** shows the partial regression of the MSSS score on IgA-GLIAL-CAM-370-399 (after adjusting for the effects of age and sex).

**Table 4.**
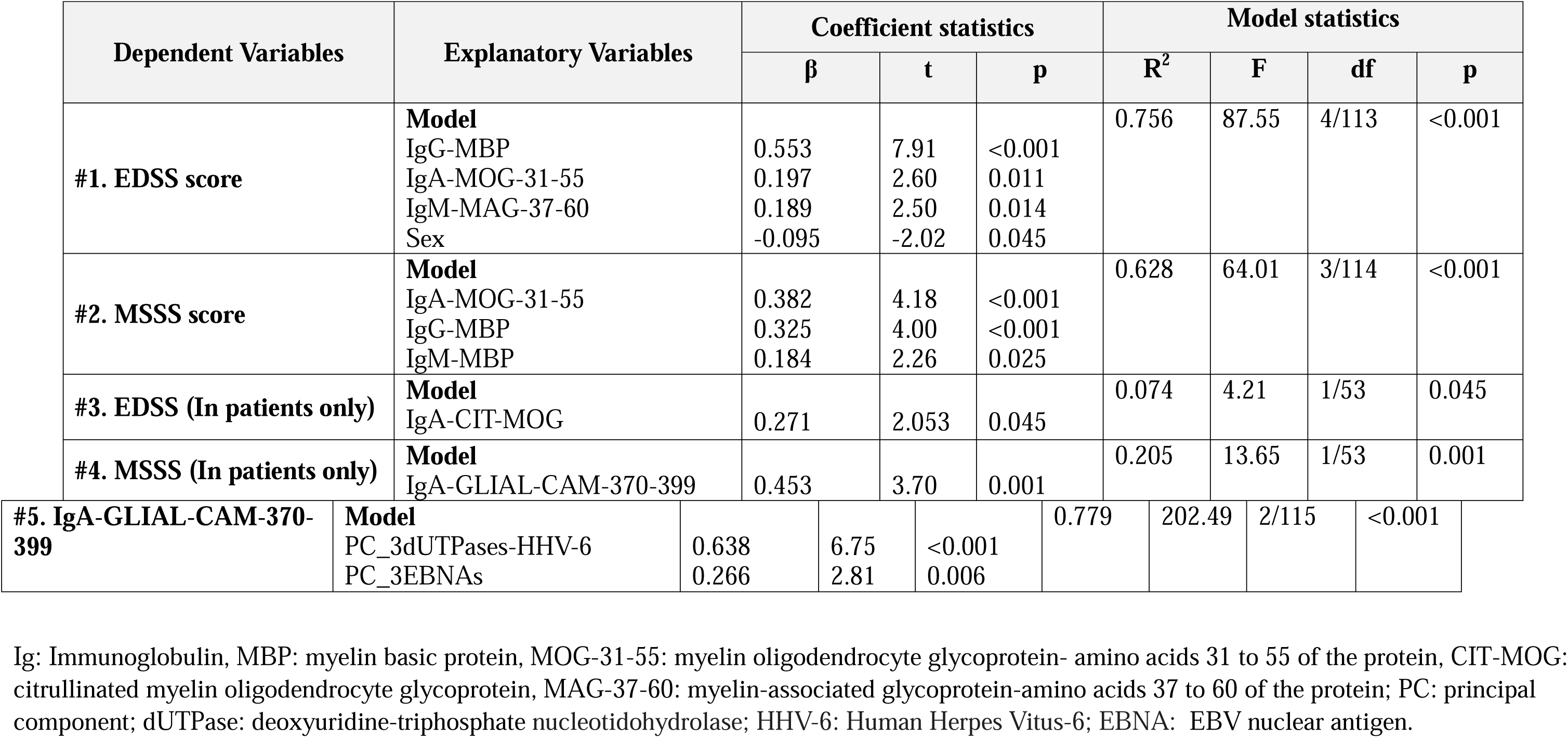
Results of multiple regression analysis with the Expanded Disability Status Scale (EDSS) and Multiple Sclerosis Severity Score (MSSS) ratings and IgA directed against GLIAL-CAM as dependent variables.

**Figure 4:**
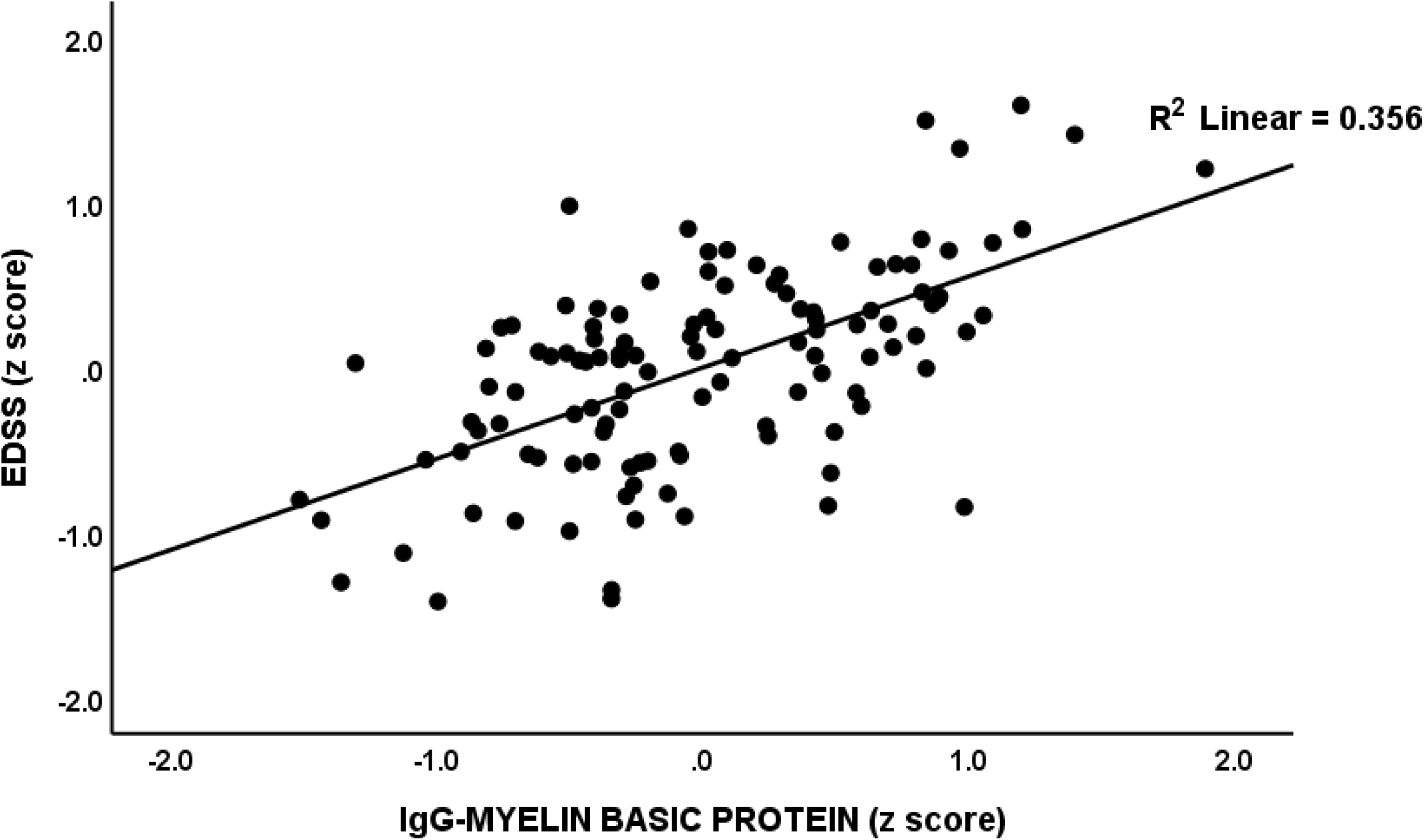
Partial regression of the Expanded Disability Status Scale (EDSS) on immunoglobulin (Ig)G directed against myelin basic protein, p < 0.001.

**Figure 5:**
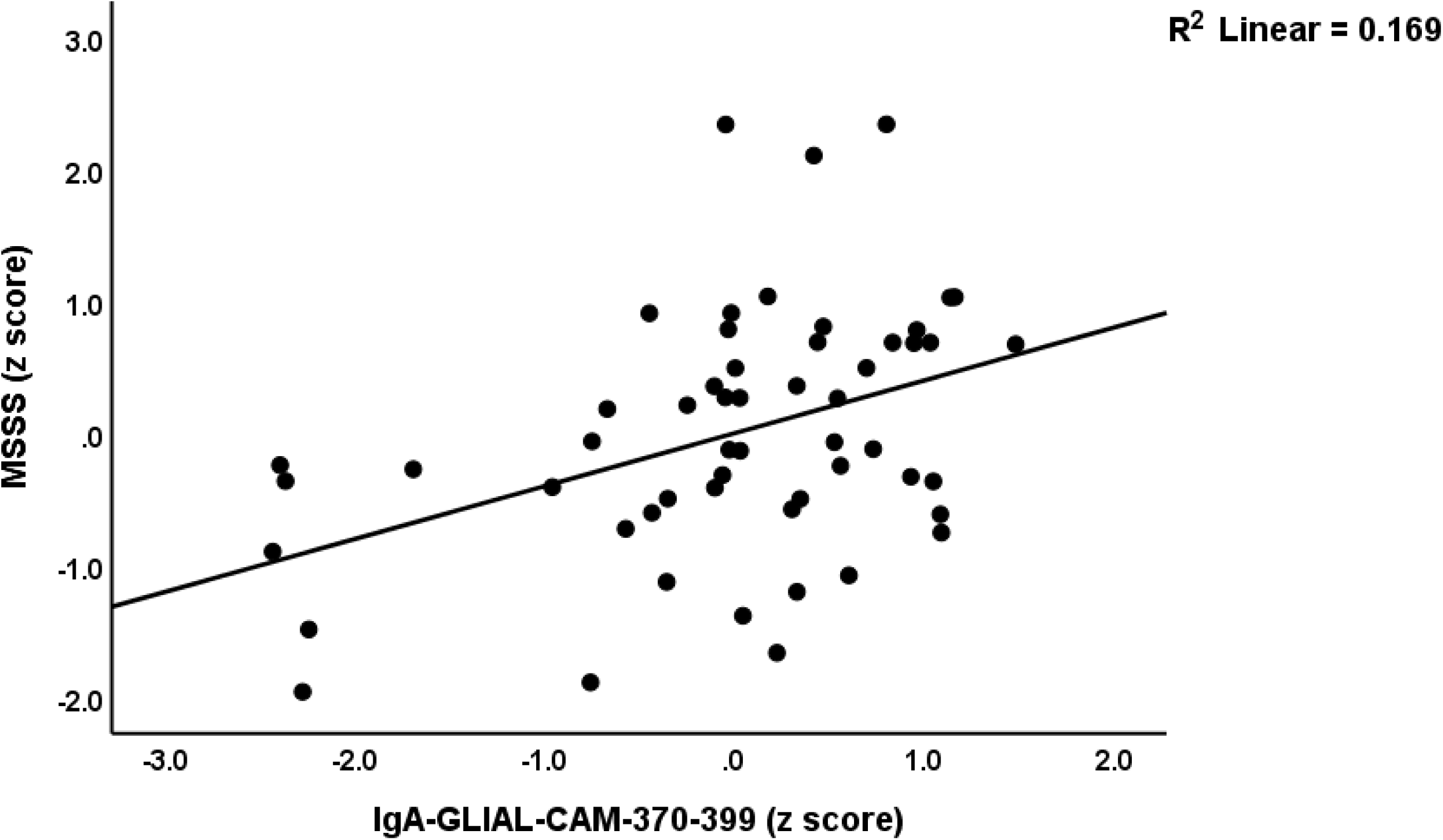
Partial regression of the multiple sclerosis severity score (MSSS) on immunoglobulin (Ig)A directed against GLIAL-CAM, p < 0.001.

We have also examined, in the restricted group of patients, the correlations between the EDSS and MSSS scores and the myelin-related proteins as shown in ESF, Table 2. This table also shows the associations with duration of illness (DOI). We found that the MSSS and duration of illness were significantly associated with IgM and IgA against MBP and MOG-35-55, as well as IgA responses to PLP, MOG-35-55, CIT-MOG, MAG-37-60, and GLIAL-CAM-370-399. The EDSS scores were found to be positively correlated with IgA levels to MOG-35-55 and CIT-MOG. The EDSS and MSSS score were significantly and positively associated with the autoimmune responses, whereas DOI was negatively associated with the markers.

### Viral replication and autoimmune responses to myelin-related proteins

To examine the relationship between immune responses to viral antigens and the autoimmune reactions targeting myelin-associated proteins, canonical correlation analysis was used. The autoimmune response to myelin served as the dependent variable in this analysis. First, we computed the first PCs from the immunoglobulin responses to all myelin-associated markers (labeled PC_IgGallmyelin, PC_IgAallmyelin, and PC_IgMallmyelin) and introduced these as dependent variables (set 1). Second, as explained above, we computed PC_3EBNAs, PC_3dUTPases-EBV, and PC_3dUTPases-HHV-6 and used these as explanatory variables (set 2). As shown in **Table 5**, significant canonical components could be derived from both the immune responses to myelin-related proteins and viral antigens. Moreover, 47.4% of the variance in set 1 (the myelin-related proteins) was predicted by set 2 (the viral antigens).

**Table 5.**
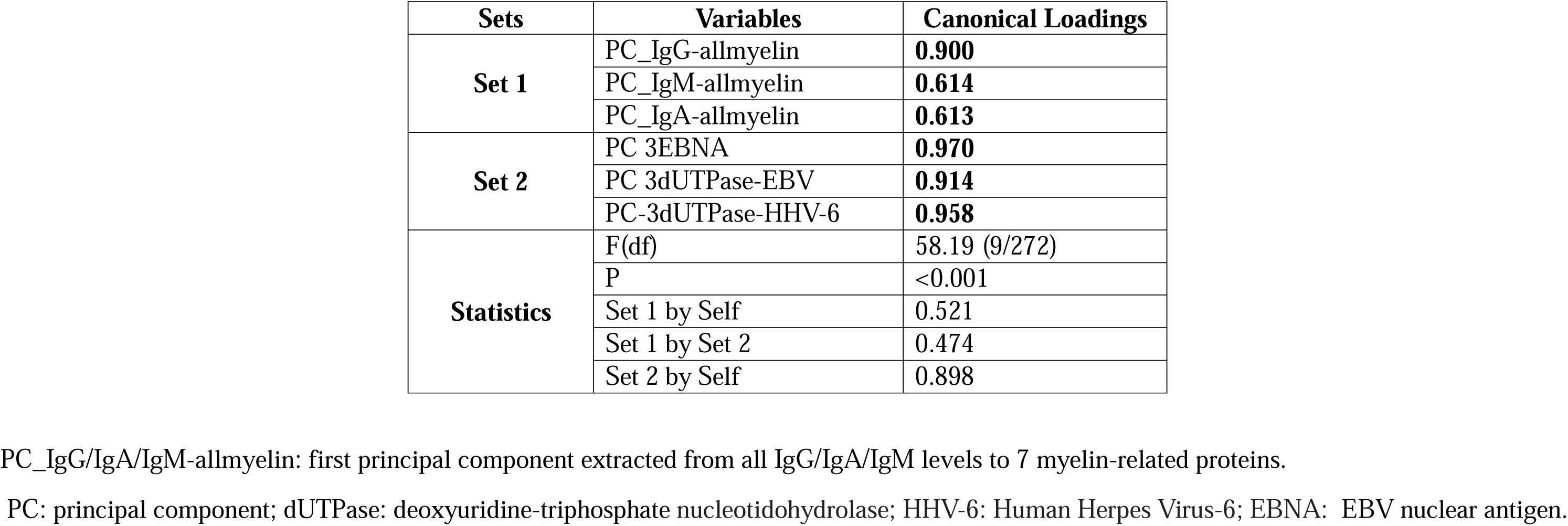
Results of canonical correlation analysis with autoimmune biomarkers as dependent variables and immune reactivity to latent viruses as explanatory variables.

Since it is known that EBNA1 may cross-react with GLIAL-CAM [30] we have examined whether IgG/IgA/IgM responsivity to EBNA is a significant predictor of IgA-GLIAL-CAM (the most important GLIAL-CAM biomarker in the same table, see regression #4). Table 4, regression #5 shows the outcome of an automatic multivariate regression analysis with IgA-GLIAL-CAM (on the immune responses to viral antigens while allowing for the effects of age, sex, BMI, etc. We found that 77.9% of the variance in IgA-GLIAL-CAM was explained by PC_3EBNAs and PC_3UTPAases-HHV-6. Figure 6 shows the partial regression of IgA-GLIAL-CAM on IgA-EBNA.

**Figure 6:**
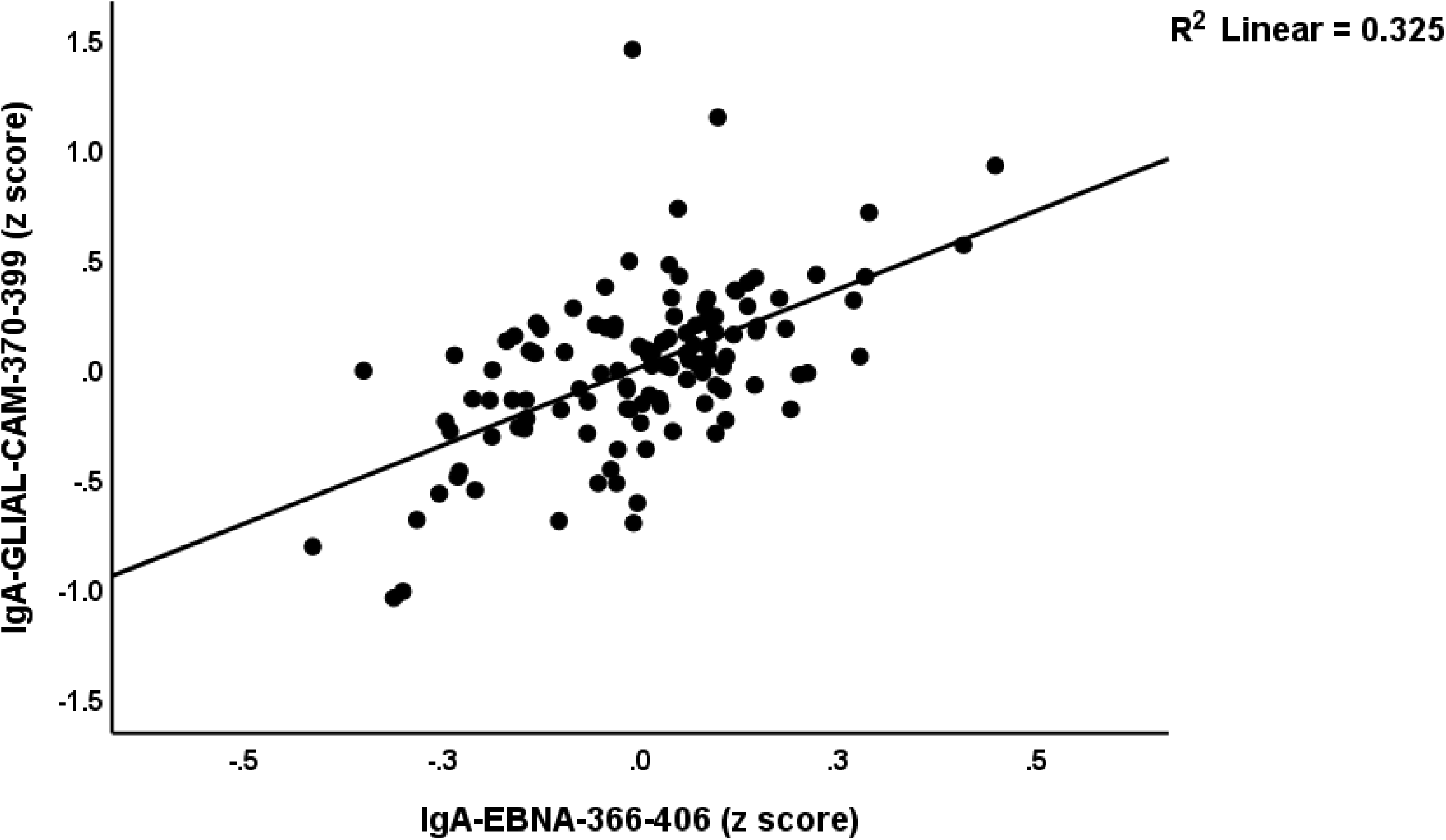
Partial renucleargression of immunoglobulin (Ig)A directed to GLIAL-CAM on immunoglobulin (Ig)A directed to Epstein-Barr Virus antigen (EBNA)-366-406, p < 0.001.

## Discussion

### Autoantibodies targeting key myelin proteins in RRMS

Our study’s primary finding reveals a distinctive autoimmune profile in patients with RRMS, characterized by significantly elevated IgA, IgM, and IgG antibodies levels targeting key myelin proteins. These include MBP, PLP, MOG-35-55, MOG-31-55, CIT-MOG), MAG-37-60, and glialCAM-370-399. These findings suggest a significant autoimmune response against myelin-associated proteins, contributing to the disease process in RRMS. Importantly, our study reveals that the IgG reaction to MBP and the IgM response to MAG-37-60 may differentiate RRMS patients from healthy controls with 96.6% accuracy. Moreover, neural network analysis showed that 4 markers significantly discriminated RRMS from controls with a predictive accuracy of 95.5% (in a holdout sample) with sensitivity=95.7% and specificity of 95.2%. Apart from IgG levels directed to MBP, also IgA levels directed to MBP, IgM to MAG-37-60, and IgA to MOG-31-55 were highly significant discriminators. As such, the diagnostic performance of IgG directed to MBP may be improved by adding IgA and IgM reactivity to these myelin-related proteins.

Consistent with our findings, previous work has shown that IgM autoantibodies targeting myelin proteins like MBP, PLP, and MOG are elevated in RRMS patients [20, 31]. However, earlier attempts to use MOG antibodies as MS biomarkers were less successful [32], highlighting the unique importance of the IgA and IgM antibodies in our study. Other autoantibodies, such as IgM responses to phosphatidylcholine and antibodies directed to MBP and MAG found within extracellular vesicles, have been proposed as potential biomarkers in RRMS [33, 34]. These findings suggest a broader autoimmune response targeting multiple CNS proteins in RRMS. Future research should explore whether these markers can distinguish RRMS from other MS subtypes, such as primary progressive MS.

### Autoimmune Responses to myelin-related proteins, EDSS and MSSS

The second major finding of our study indicates that elevated concentrations of autoantibodies directed against the major myelin proteins, including IgG/IgM-MBP, IgA-MOG-31-55, and IgM-MAG-37-60, have cumulative effects in predicting increased disability and disease severity scores. Moreover, in patients with RRMS, we found that increases in IgA-CIT-MOG predicted the EDSS score, and that IgA-GLIAL-CAM-370-399 predicted the MSSS score. Thus, the severity and progression of disease are not only associated with MBP antibody demyelination but also to MOG and MAG antibody demyelination, and damage to GLIAL-CAM and to a lesser degree also PLP. These findings underscore that wider autoimmune reactions targeting multiple myelin self-antigens exacerbate disease severity and progression in RRMS.

Our study extends the findings of previous reports demonstrating that autoantibodies targeting myelin-related constituents, including MBP and MOG, are associated with heightened disease activity and severity in MS disease [35, 36]. For example, increased CSF concentrations of these immunoglobulins predict more severe disease trajectories, including reduced intervals to EDSS deterioration [37, 38]. This aligns with the concept that intrathecal IgM and IgG synthesis significantly contributes to inflammation and demyelination, resulting in neurological deterioration [39]. Moreover, increased CSF IgA and IgM concentrations have been associated with cortical atrophy and functional decline in RRMS patients [40], underscoring the pathogenic relevance of these antibodies in disease progression. The proteolytic activity of these antibodies towards MBP has been suggested as a direct factor in MS pathogenesis [41], emphasizing the active involvement of autoantibodies in the neurodegenerative mechanisms of RRMS.

### Viral reactivation and autoantibodies targeting key myelin proteins

The third key finding of our study reveals a strong association between the reactivation of both viruses (as measured with IgG/IgA/IgM responses towards dUTPases of both viruses and towards EBNA) [23] and autoimmune responses targeting the essential myelin proteins. Thus, the viral reactivation markers are not only associated with increased disease severity and disease progression [23] but also may constitute key drivers of the autoimmune responses to myelin-related proteins. This suggests that replication of HHV-6 and EBV may lead to CNS demyelination and the symptoms and progression of RRMS.

Previous studies have supported the link between viral reactivation and autoimmune responses in diseases like Long COVID, where persistence of SARS-CoV-2 and reactivation of HHV-6 were significantly correlated with autoimmune responses targeting CNS proteins such as MBP, MOG, and synapsin [29]. This reinforces the role of viral persistence and reactivation in triggering autoimmunity.

The association between latent viruses’ reactivation and (EBV and HHV6) and MS has been well-documented [23]. EBV infects B-lymphocytes, which can migrate to the CNS, forming inflammatory aggregates that contribute to neuroinflammation and demyelination [42]. These aggregates consist of macrophages, astrocytes, and lymphocytes, which further exacerbate neurodegenerative processes. Antibodies against EBV antigens, particularly Epstein-Barr Nuclear Antigen 1 (EBNA-1), have been shown to cross-react with myelin proteins, including MBP, through molecular mimicry, leading to myelin destruction [43]. The concept of molecular mimicry between viral antigens and myelin proteins may be central to the pathogenesis of MS. In this respect, it has been suggested that antibodies against EBV’s latent membrane protein 1 (LMP1) may cross-react with MBP, triggering an autoimmune response that targets myelin [44]. Likewise, there is evidence of high-affinity molecular mimicry between EBNA1 and GIAL-CAM, suggesting that antibodies initially targeting EBNA1 due to EBV infection may cross-react with GLIAL-CAM, thereby contributing to RRMS pathology [30]. This association is further corroborated by our findings that a large part of the variance in the most important GLIAL-CAM biomarker (namely IgA) is strongly predicted by the immune responsivity to EBNA and HHV-6.

In this regard, HHV-6 can infect glial precursor cells, altering their morphology and impairing their ability to repair myelin [45]. In experimental models, HHV-6 has been shown to induce apoptosis in CNS cells, contributing to chronic demyelination and increased T-cell reactivity to myelin antigens [46]. Additionally, HHV-6 infection promotes the release of inflammatory cytokines such as IL-6 and chemokines like CXCL13, which further aggravate neuroinflammation and contribute to the demyelination process [47].

Moreover, chronic exposure to viral antigens may lead to epitope spreading, where the immune response that initially targets viral proteins expands to include other myelin-related proteins [44]. This immune expansion may contribute to the progressive nature of RRMS and the continued attack on many myelin-related components. Overall, the reactivation of these latent viruses not only increases neuroinflammation but also perpetuates the autoimmune attack on myelin, highlighting their role in the demyelination process observed in RRMS.

## Limitations

The present findings should be interpreted considering several limitations. First, this study focused exclusively on patients with RRMS; thus, it would be essential to assess these CNS autoimmune biomarkers in other MS subtypes, such as primary progressive MS and secondary progressive MS, to determine whether similar patterns are present across the spectrum of the disease. It would have been even more interesting if we had included oxidative and nitrosative stress biomarkers [8, 48]. Incorporating these biomarkers in future studies would provide a more comprehensive understanding of how oxidative stress interacts with autoimmune responses leading to damage to myelin-associated proteins. Finally, our study was performed in patients who were in a remitted phase of RRMS. Therefore, future research should investigate the associations among clinical symptoms, autoimmunity, and viral reactivation in RRMS patients during acute relapses as compared with the remitted phase. Future research should confirm the role of the autoimmunity biomarkers of RRMS established in the current study in other countries and ethnicities.

## Conclusion

This study demonstrates that RRMS is associated with a significant elevation of IgG/IgA/IgM autoimmune responses targeting multiple key myelin-related proteins. The findings suggest that certain autoantibodies may have potential as both diagnostic and prognostic biomarkers for RRMS, aiding in the identification of disease activity and progression. Moreover, the reactivation of latent viruses, such as EBV and HHV-6, appears to play a critical role in triggering or maintaining autoimmune responses in RRMS. The findings highlight the necessity of monitoring viral reactivation in patients with RRMS, as it may influence disease severity and progression. Future research should investigate the impact of antiviral treatments on autoimmune responses to myelin-related proteins in RRMS.

## Data Availability

The corresponding author (MM) is available to provide access to the dataset related to this study upon receiving a valid request and following a thorough review of the data.

## Acknowledgements

The authors are deeply grateful to the Neuroscience Center of Alsader Medical City in Al-Najaf province, Iraq, for their invaluable support in the data collection process.

## Ethical approval and consent to participate

The Ethics Committee of the College of Medical Technology at the Islamic University of Najaf, Iraq, granted sanction for the investigation (Document No. 11/2021). Written informed consent was obtained from all patients and control participants, and all procedures were conducted in accordance with Iraqi and international ethical standards.

## Author’s contributions

AFA supervised the blood sample collection and patient procedures. Serum biomarker quantification was performed by AV and EV at Immunosciences Lab, who assumed the costs of testing. MM conducted the statistical analysis. MM and AV created the visualizations. AFA drafted the initial manuscript, which was reviewed and revised by MM, AV, EV, JL and YZ. All authors approved the last version of the manuscript.

## ELECTRONIC SUPPLEMENTARY FILE (ESF)

ESF, Table 1. Sociodemographic and clinical data in patients with relapsing-remitting multiple sclerosis (RRMS) and healthy controls (HC).

| **Variables** | **Healthy Controls (n=63)** | **RRMS Patients (n=55)** | **F/$\chi^2$** | **df** | **p-value** |
| --- | --- | --- | --- | --- | --- |
| Age (years) | 31.4 (7.1) | 29.5 (6.3) | 2.18 | 1/116 | 0.142 |
| Sex (M/F) | 33/30 | 37/18 | 2.70 | 1 | 0.133 |
| BMI (kg/m2) | 26.33 (4.50) | 24.94 (3.52) | 3.41 | 1/116 | 0.068 |
| Education (years) | 17.2 (5.1) | 18.4 (3.4) | 2.04 | 1/116 | 0.155 |
| Married/Separated | 26/37 | 31/24 | 2.68 | 1 | 0.139 |
| Smoking (N/Y) | 44/19 | 50/5 | 8.04 | 1 | 0.006 |
| Employment (N/Y) | 2/28 | 46/9 | 46.78 | 1 | <0.001 |
| MSSS | 0 | 1.71 (1.20) | MWU | - | <0.001 |
| EDSS | 0 | 1.03 (0.11) | MWU | - | <0.001 |
| PC-Severity | -0.921(0) | 1.05(0.198) | MWU | 1/116 | <0.001 |
| PC_3dUTPase-EBV | -0.662(0.745) | 0.759(0.655) | 119.51 | 1/116 | <0.001 |
| PC_3dUTPase-HHV-6 | -0.697(0.694) | 0.798(0.628) | 148.71 | 1/116 | <0.001 |
| PC_3EBNA | -0.579(0.810) | 0.664(0.755) | 73.71 | 1/116 | <0.001 |

M: Male, F: Female, kg: Kilogram, m2: squared meter, N: No, Y: Yes, MSSS: Multiple Sclerosis Severity Score, EDSS: Expanded Disability Status Scale, PC-Severity: first principal component extracted from EDSS and MSSS, PC 3dUTPase-EBV: first principal component extracted from IgA, IgM,IgG directed to Epstein-Barr Virus deoxyuridine-triphosphatase (dUTPase-EBV), PC 3dUTPase-HHV-6: first principal component extracted from IgA, IgM,IgG directed to herpesvirus 6 deoxyuridine-triphosphatase (dUTPase-HHV-6), PC 6dUTPases: first principal component extracted from IgA, IgM,IgG directed to both dUTPase-EBV and dUTPase-HHV-6, PC 3EBNA: first principal component extracted from IgA, IgM,IgG directed to Epstein-Barr virus nuclear antigen 1 (EBNA).

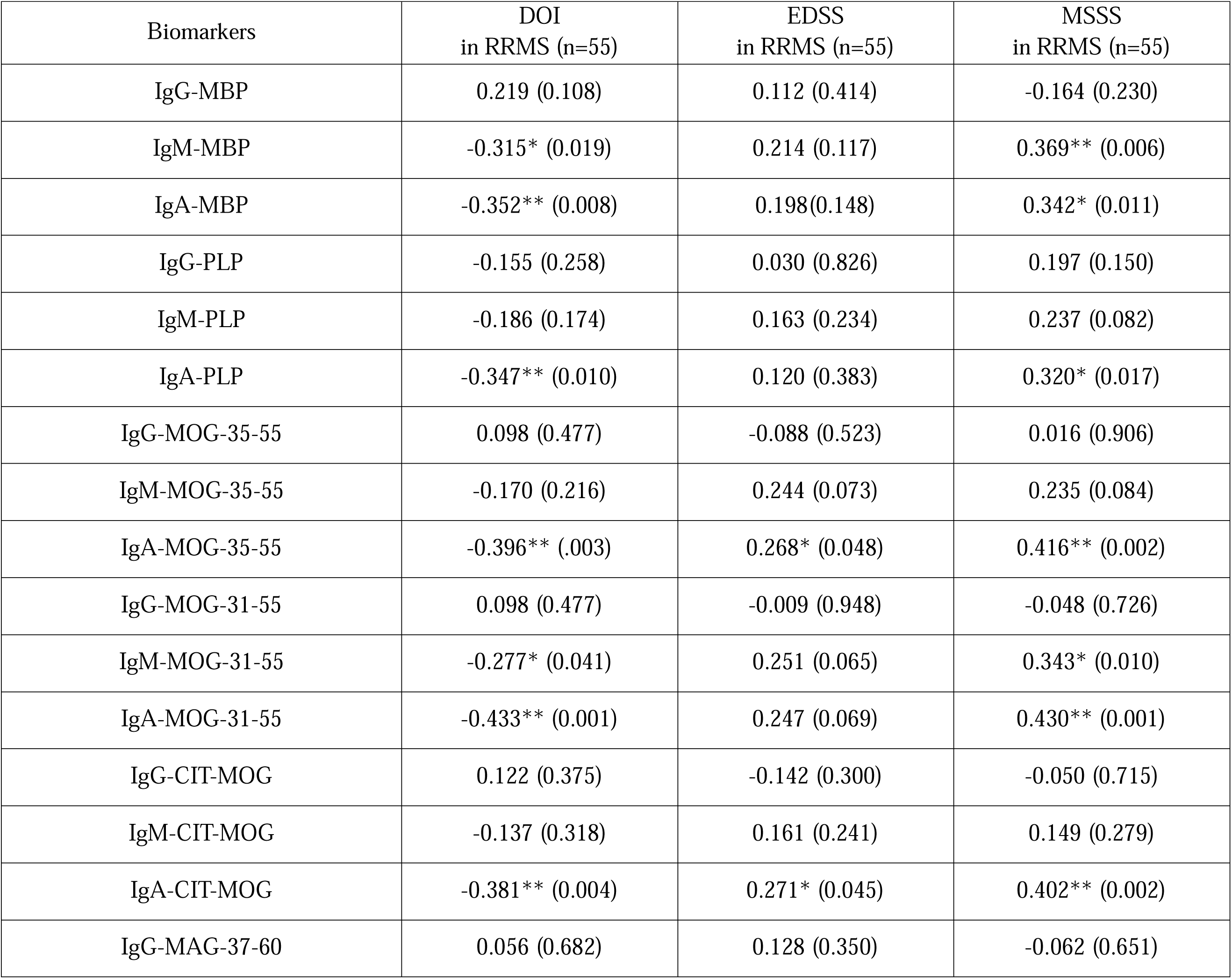
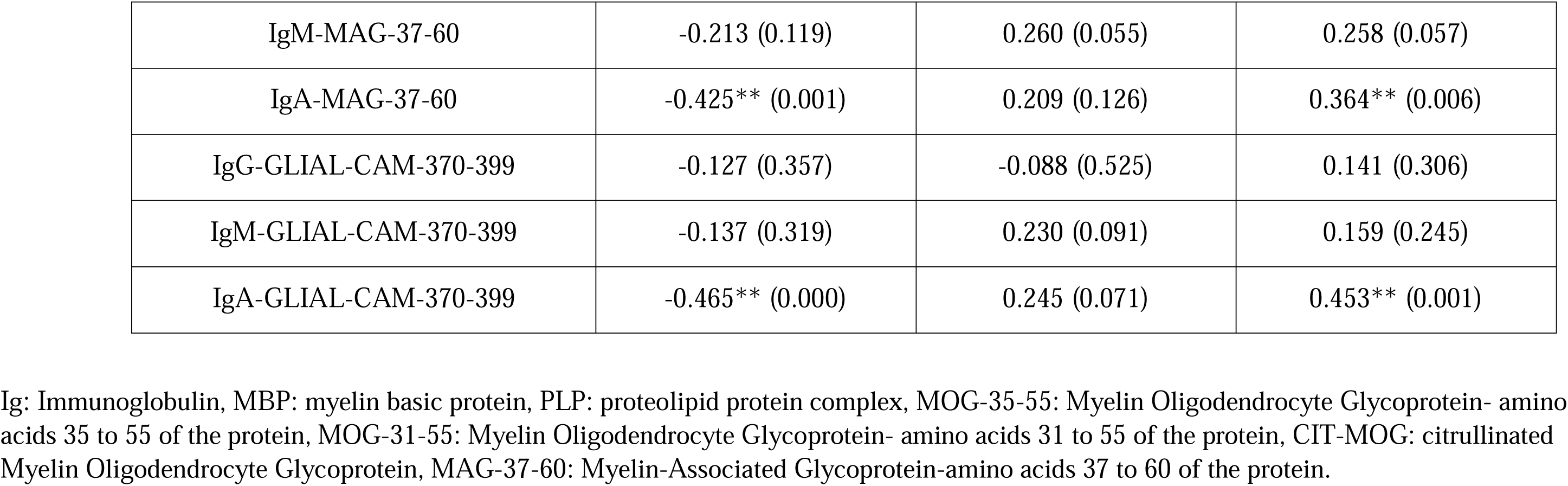
ESF, Table 2. Intercorrelation matrix between between IgG, IgA and IgM against viral antigens and neuropsychiatric rating scales in relapsing-remitting multiple sclerosis (RRMS).

| Biomarkers | DOI<br>in RRMS (n=55) | EDSS<br>in RRMS (n=55) | MSSS<br>in RRMS (n=55) |
| --- | --- | --- | --- |
| IgG-MBP | 0.219 (0.108) | 0.112 (0.414) | -0.164 (0.230) |
| IgM-MBP | -0.315* (0.019) | 0.214 (0.117) | 0.369** (0.006) |
| IgA-MBP | -0.352** (0.008) | 0.198(0.148) | 0.342* (0.011) |
| IgG-PLP | -0.155 (0.258) | 0.030 (0.826) | 0.197 (0.150) |
| IgM-PLP | -0.186 (0.174) | 0.163 (0.234) | 0.237 (0.082) |
| IgA-PLP | -0.347** (0.010) | 0.120 (0.383) | 0.320* (0.017) |
| IgG-MOG-35-55 | 0.098 (0.477) | -0.088 (0.523) | 0.016 (0.906) |
| IgM-MOG-35-55 | -0.170 (0.216) | 0.244 (0.073) | 0.235 (0.084) |
| IgA-MOG-35-55 | -0.396** (.003) | 0.268* (0.048) | 0.416** (0.002) |
| IgG-MOG-31-55 | 0.098 (0.477) | -0.009 (0.948) | -0.048 (0.726) |
| IgM-MOG-31-55 | -0.277* (0.041) | 0.251 (0.065) | 0.343* (0.010) |
| IgA-MOG-31-55 | -0.433** (0.001) | 0.247 (0.069) | 0.430** (0.001) |
| IgG-CIT-MOG | 0.122 (0.375) | -0.142 (0.300) | -0.050 (0.715) |
| IgM-CIT-MOG | -0.137 (0.318) | 0.161 (0.241) | 0.149 (0.279) |
| IgA-CIT-MOG | -0.381** (0.004) | 0.271* (0.045) | 0.402** (0.002) |
| IgG-MAG-37-60 | 0.056 (0.682) | 0.128 (0.350) | -0.062 (0.651) |
| IgM-MAG-37-60 | -0.213 (0.119) | 0.260 (0.055) | 0.258 (0.057) |
| IgA-MAG-37-60 | -0.425** (0.001) | 0.209 (0.126) | 0.364** (0.006) |
| IgG-GLIAL-CAM-370-399 | -0.127 (0.357) | -0.088 (0.525) | 0.141 (0.306) |
| IgM-GLIAL-CAM-370-399 | -0.137 (0.319) | 0.230 (0.091) | 0.159 (0.245) |
| IgA-GLIAL-CAM-370-399 | -0.465** (0.000) | 0.245 (0.071) | 0.453** (0.001) |

Ig: Immunoglobulin, MBP: myelin basic protein, PLP: proteolipid protein complex, MOG-35-55: Myelin Oligodendrocyte Glycoprotein- amino acids 35 to 55 of the protein, MOG-31-55: Myelin Oligodendrocyte Glycoprotein- amino acids 31 to 55 of the protein, CIT-MOG: citrullinated Myelin Oligodendrocyte Glycoprotein, MAG-37-60: Myelin-Associated Glycoprotein-amino acids 37 to 60 of the protein.

## References

[1] M. Niino, Y. Miyazaki. [Relapsing-Remitting Multiple Sclerosis]. Brain Nerve, 2021;73:442–9.

[2] S. Goonesekera. A Global Epidemiological Forecast of Multiple Sclerosis and Disease Subtypes (S16.002). Neurology, 2017;88:S16.002.

[3] F. D. Lublin, S. C. Reingold. Defining the clinical course of multiple sclerosis: results of an international survey. National Multiple Sclerosis Society (USA) Advisory Committee on Clinical Trials of New Agents in Multiple Sclerosis. Neurology, 1996;46:907–11.

[4] H. Lassmann. Multiple Sclerosis Pathology. Cold Spring Harb Perspect Med, 2018;8.

[5] P. Villoslada, S. Llufriu. Preventing Neurodegeneration in Multiple Sclerosis Is Required From the Earliest Stages of the Disease. Neurology, 2022;99:641–2.

[6] B. D. Trapp, R. Ransohoff, R. Rudick. Axonal pathology in multiple sclerosis: relationship to neurologic disability. Curr Opin Neurol, 1999;12:295–302.

[7] B. Nourbakhsh, E. M. Mowry. Multiple Sclerosis Risk Factors and Pathogenesis. Continuum (Minneap Minn), 2019;25:596–610.

[8] A. P. Kallaur, E. M. V. Reiche, S. R. Oliveira, A. N. C. Simão, W. L. d. C. J. Pereira, D. F. Alfieri et al. Genetic, Immune-Inflammatory, and Oxidative Stress Biomarkers as Predictors for Disability and Disease Progression in Multiple Sclerosis. Molecular Neurobiology, 2017;54:31–44.

[9] L. Mosca, V. Mantero, S. Penco, L. La Mantia, S. De Benedetti, M. R. Marazzi et al. HLA-DRB1*15 association with multiple sclerosis is confirmed in a multigenerational Italian family. Funct Neurol, 2017;32:83–8.

[10] M. B. Sintzel, M. Rametta, A. T. Reder. Vitamin D and Multiple Sclerosis: A Comprehensive Review. Neurol Ther, 2018;7:59–85.

[11] S. Hempel, G. D. Graham, N. Fu, E. Estrada, A. Y. Chen, I. Miake-Lye et al. A systematic review of modifiable risk factors in the progression of multiple sclerosis. Mult Scler, 2017;23:525–33.

[12] Z. Khalesi, V. Tamrchi, M. H. Razizadeh, A. Letafati, P. Moradi, A. Habibi et al. Association between human herpesviruses and multiple sclerosis: A systematic review and meta-analysis. Microbial Pathogenesis, 2023;177:106031.

[13] A. F. Almulla, A. K. Abdul Jaleel, A. A. Abo Algon, C. Tunvirachaisakul, H. K. Hassoun, H. K. Al-Hakeim et al. Mood Symptoms and Chronic Fatigue Syndrome Due to Relapsing-Remitting Multiple Sclerosis Are Associated with Immune Activation and Aberrations in the Erythron. Brain Sci, 2023;13.

[14] A. P. Kallaur, S. R. Oliveira, A. N. C. Simão, D. F. Alfieri, T. Flauzino, J. Lopes et al. Cytokine Profile in Patients with Progressive Multiple Sclerosis and Its Association with Disease Progression and Disability. Mol Neurobiol, 2017;54:2950–60.

[15] A. P. Kallaur, J. Lopes, S. R. Oliveira, A. N. Simão, E. M. Reiche, E. R. de Almeida et al. Immune-Inflammatory and Oxidative and Nitrosative Stress Biomarkers of Depression Symptoms in Subjects with Multiple Sclerosis: Increased Peripheral Inflammation but Less Acute Neuroinflammation. Mol Neurobiol, 2016;53:5191–202.

[16] N. Kaushansky, M. Eisenstein, R. Zilkha-Falb, A. Ben-Nun. The myelin-associated oligodendrocytic basic protein (MOBP) as a relevant primary target autoantigen in multiple sclerosis. Autoimmun Rev, 2010;9:233–6.

[17] N. K. de Rosbo, A. Ben-Nun. T-cell Responses to Myelin Antigens in Multiple Sclerosis; Relevance of the Predominant Autoimmune Reactivity to Myelin Oligodendrocyte Glycoprotein. Journal of Autoimmunity, 1998;11:287–99.

[18] P. Stinissen, J. C. M. Raus. Autoreactive T lymphocytes in multiple sclerosis: pathogenic role and therapeutic targeting. Acta neurologica Belgica, 1999;99 1:65–9.

[19] A. Ganesan, P. Muralidharan, L. N. Ramya. The Fulcrum of Demyelination in Multiple Sclerosis. Curr Protein Pept Sci, 2023;24:579–88.

[20] A. Vojdani, E. Vojdani, E. Cooper. Antibodies to myelin basic protein, myelin oligodendrocytes peptides, alpha-beta-crystallin, lymphocyte activation and cytokine production in patients with multiple sclerosis. J Intern Med, 2003;254:363–74.

[21] U. C. Meier, R. C. Cipian, A. Karimi, R. Ramasamy, J. M. Middeldorp. Cumulative Roles for Epstein-Barr Virus, Human Endogenous Retroviruses, and Human Herpes Virus-6 in Driving an Inflammatory Cascade Underlying MS Pathogenesis. Front Immunol, 2021;12:757302.

[22] R. E. Tarlinton, E. Martynova, A. A. Rizvanov, S. Khaiboullina, S. Verma. Role of Viruses in the Pathogenesis of Multiple Sclerosis. Viruses, 2020;12.

[23] A. F. Almulla, A. Vojdani, Y. Zhang, E. Vojdani, M. Maes. Reactivation of Human Herpesvirus 6 and Epstein-Barr Virus in relapsing remitting multiple sclerosis: association with disabilities, disease progression, and inflammatory processes. medRxiv, 2024:2024.09.10.24313388.

[24] M. Maes, A. F. Almulla, E. Vojdani, E. Dzhambazova, D. Stoyanov, Y. Zhang et al. Chronic fatigue syndrome, depression, and anxiety symptoms due to relapsing-remitting multiple sclerosis are associated with reactivation of Epstein-Barr virus and Human Herpesvirus 6. medRxiv, 2024:2024.10.12.24315393.

[25] C. H. Polman, S. C. Reingold, B. Banwell, M. Clanet, J. A. Cohen, M. Filippi et al. Diagnostic criteria for multiple sclerosis: 2010 revisions to the McDonald criteria. Annals of neurology, 2011;69:292–302.

[26] J. F. Kurtzke. Rating neurologic impairment in multiple sclerosis: an expanded disability status scale (EDSS). Neurology, 1983;33:1444-.

[27] R. Roxburgh, S. Seaman, T. Masterman, A. Hensiek, S. Sawcer, S. Vukusic et al. Multiple Sclerosis Severity Score: using disability and disease duration to rate disease severity. Neurology, 2005;64:1144–51.

[28] N. H. Trier, B. E. Holm, J. Heiden, O. Slot, H. Locht, H. Lindegaard et al. Antibodies to a strain-specific citrullinated Epstein-Barr virus peptide diagnoses rheumatoid arthritis. Scientific Reports, 2018;8:3684.

[29] A. F. Almulla, M. Maes, B. Zhou, H. K. Al-Hakeim, A. Vojdani. Brain-targeted autoimmunity is strongly associated with Long COVID and its chronic fatigue syndrome as well as its affective symptoms. medRxiv, 2023:2023.10.04.23296554.

[30] T. V. Lanz, R. C. Brewer, P. P. Ho, J. S. Moon, K. M. Jude, D. Fernandez et al. Clonally expanded B cells in multiple sclerosis bind EBV EBNA1 and GlialCAM. Nature, 2022;603:321–7.

[31] K. Van Haren, B. H. Tomooka, B. A. Kidd, B. Banwell, A. Bar-Or, T. Chitnis et al. Serum autoantibodies to myelin peptides distinguish acute disseminated encephalomyelitis from relapsing-Remitting multiple sclerosis. Multiple Sclerosis Journal, 2013;19:1726–33.

[32] S. Amor, G. Giovannoni. Antibodies to myelin oligodendrocyte glycoprotein as a biomarker in multiple sclerosis--are we there yet? Mult Scler, 2007;13:1083–5.

[33] M. C. Sádaba, V. Rothhammer, Ú. Muñoz, C. Sebal, E. Escudero, P. Kivisäkk et al. Serum antibodies to phosphatidylcholine in MS. Neurology: Neuroimmunology and NeuroInflammation, 2020;7.

[34] C. Agliardi, F. R. Guerini, M. Zanzottera, E. Bolognesi, S. Picciolini, D. Caputo et al. Myelin Basic Protein in Oligodendrocyte-Derived Extracellular Vesicles as a Diagnostic and Prognostic Biomarker in Multiple Sclerosis: A Pilot Study. International Journal of Molecular Sciences, 2023;24.

[35] R. Egg, M. Reindl, F. Deisenhammer, C. Linington, T. Berger. Anti-MOG and anti-MBP antibody subclasses in multiple sclerosis. Mult Scler, 2001;7:285–9.

[36] F. Angelucci, M. Mirabella, G. Frisullo, M. Caggiula, P. A. Tonali, A. P. Batocchi. Serum levels of anti-myelin antibodies in relapsing-remitting multiple sclerosis patients during different phases of disease activity and immunomodulatory therapy. Dis Markers, 2005;21:49–55.

[37] P. Perini, F. Ranzato, M. Calabrese, L. Battistin, P. Gallo. Intrathecal IgM production at clinical onset correlates with a more severe disease course in multiple sclerosis. Journal of Neurology, Neurosurgery & Psychiatry, 2006;77:953.

[38] I. Rosenstein, S. Rasch, M. Axelsson, L. Novakova, K. Blennow, H. Zetterberg et al. Increased intrathecal neurofilament light and immunoglobulin M predict severe disability in relapsing-remitting multiple sclerosis. Frontiers in Immunology, 2022;13.

[39] C. Gasperi, A. Salmen, G. Antony, A. Bayas, C. Heesen, T. Kümpfel et al. Association of Intrathecal Immunoglobulin G Synthesis With Disability Worsening in Multiple Sclerosis. JAMA Neurology, 2019;76:841–9.

[40] J. Kroth, D. Ciolac, V. Fleischer, N. Koirala, J. Krämer, M. Muthuraman et al. Increased cerebrospinal fluid albumin and immunoglobulin A fractions forecast cortical atrophy and longitudinal functional deterioration in relapsing-remitting multiple sclerosis. Mult Scler, 2019;25:338–43.

[41] D. I. Polosukhina, V. N. Buneva, B. M. Doronin, O. B. Tyshkevich, A. N. Boiko, E. I. Gusev et al. Hydrolysis of myelin basic protein by IgM and IgA antibodies from the sera of patients with multiple sclerosis. Med Sci Monit, 2005;11:Br266–72.

[42] A. Hassani, N. Reguraman, S. Shehab, G. Khan. Primary Peripheral Epstein-Barr Virus Infection Can Lead to CNS Infection and Neuroinflammation in a Rabbit Model: Implications for Multiple Sclerosis Pathogenesis. Frontiers in Immunology, 2021;12.

[43] T. O. Ojo, O. E. Elegbeleye, O. Q. Bolaji, T. I. Adelusi, E. K. Oladipo, M. O. Olawuyi et al. Hitting Epstein Barr virus where it hurts: computational methods exploration for siRNA therapy in alleviating Epstein Barr virus-induced multiple sclerosis. Neurogenetics, 2024;25:263–75.

[44] Y. Lomakin, G. P. Arapidi, A. Chernov, R. Ziganshin, E. Tcyganov, I. Lyadova et al. Exposure to the Epstein-Barr Viral Antigen Latent Membrane Protein 1 Induces Myelin-Reactive Antibodies In Vivo. Front Immunol, 2017;8:777.

[45] J. Dietrich, B. M. Blumberg, M. Roshal, J. V. Baker, S. D. Hurley, M. Mayer-Pröschel et al. Infection with an endemic human herpesvirus disrupts critical glial precursor cell properties. J Neurosci, 2004;24:4875–83.

[46] C. P. Genain. Animal Models. Perspectives in Medical Virology, 2006;12:305–21.

[47] M. A. Romeo, M. S. Gilardini Montani, R. Benedetti, A. Arena, A. Gaeta, M. Cirone. The dysregulation of autophagy and ER stress induced by HHV-6A infection activates pro-inflammatory pathways and promotes the release of inflammatory cytokines and cathepsin S by CNS cells. Virus Research, 2022;313.

[48] L. Mezzaroba, A. N. C. Simão, S. R. Oliveira, T. Flauzino, D. F. Alfieri, W. L. de Carvalho Jennings Pereira et al. Antioxidant and Anti-inflammatory Diagnostic Biomarkers in Multiple Sclerosis: A Machine Learning Study. Molecular Neurobiology, 2020;57:2167–78.

